# Assessment of Zero-Shot Large Language Model (LLM) Assisted Clinical Trial Matching Processes: A Metastatic Cancer Use Case

**DOI:** 10.64898/2026.07.06.26354647

**Authors:** Yingjie Weng, Himani Yalamaddi, Danning Fu, Ankita Mishra, Bryan J. Bunning, Andrew B. Martin, Jessica Hope, Vivek Charu, Allison Kurian, Manisha Desai

## Abstract

**Introduction:** For oncology patients with limited treatment options, clinical trials may be a critical lifesaving pathway. Identifying relevant trials, however, is a time-consuming and difficult task. Several patient-trial matching processes incorporating large language models (LLMs) have been proposed to alleviate the burden on patients and oncologists. We aim to explore the benefits and practical challenges of zero-shot LLM-assisted trial matching processes by analyzing the results for a single pancreatic cancer patient.

**Materials and Methods:** The results of a simple zero-shot LLM-assisted clinical trial matching process for our patient were compared to those of a “human benchmark,” which was developed manually by two of the authors interfacing directly with ClinicalTrials.gov. Performance metrics – sensitivity, specificity, precision, and accuracy – were calculated. In addition, a qualitative content analysis (QCA) of LLM reasoning text was done to identify patterns in “errors,” which we define as a human-LLM discrepancy in final patient eligibility. Implications and severity of errors are discussed.

**Results:** The zero-shot LLM-assisted process returned potential trials with a sensitivity, specificity, and precision of 81.1%, 89.3%, and 86.5% respectively compared to the human benchmark. Qualitative error analyses revealed that about 73% of errors could potentially be alleviated with improved prompting and information access. Overall performance seemed comparable to that of human reviewers.

**Conclusion:** The results from this preliminary real-world case study provide additional evidence to the literature in support of the integration of LLMs in clinical trial matching to provide benefit to patients with metastatic cancer with limited options.

## BACKGROUND AND SIGNIFICANCE

### Overview

Participation in a clinical trial for many oncology patients can be transformative and potentially lifesaving. This is especially true when an effective standard-of-care has not been established – as is often the case with complex disease or metastatic cancer – or when a first-line therapy has failed[1,2]. Despite demonstrated benefits, a 2023 estimate posited that only about 7% of adult cancer patients participated in a treatment trial[3]. Low enrollment could be attributed to the onerous nature of identifying an appropriate and accessible trial for a specific patient (which we will refer to as “patient-trial matching”)[4]. The sheer volume of available trials and their unstructured, disease-specific eligibility criteria make the task extremely time-consuming, often precluding patient-trial matching from an oncologist’s typical workflow[5]. Thus, the challenging task of locating and joining a clinical trial is often left to the patient, who likely lacks the time, resources, medical literacy, and understanding of the clinical landscape needed to determine which trials are possibilities for them[4].

### Proposed Methods

ClinicalTrials.gov[6] – an online registry of clinical research studies and results maintained by the National Library of Medicine – offers one of the most comprehensive listings of ongoing U.S. clinical trials. However, these large, ever-expanding databases provide only minimal filtering tools, and their complex interfaces and specialized medical terminology can make it challenging for patients to navigate on their own. Seeking to address the overwhelming volume of trials to review, several solutions have emerged. For example, many online patient-trial matching platforms feature questionnaires (of varying quality and detail) and sometimes employ algorithms to improve matches[7–13]. Some cancer organizations vet disease-specific trials and provide similar questionnaire-based matching tools for patients to locate them[14–16]. In our investigations, however, these questionnaires often did not sufficiently account for non-standardized patient characteristics such as molecular mutations, disease progression, or treatment histories.

LLMs, however, are well-suited to addressing these limitations, excelling at processing large amounts of unstructured, text-based information. As a result, several LLM-based systems have been proposed and implemented for both patient-to-trial matching and trial-to-patient matching (assisting clinical trialists with recruitment, which we will not address here). These systems have been quite successful regardless of the underlying LLM. However, proprietary frontier models (such as the GPT-4 series) tend to perform better – achieving strong patient-to-trial matching metrics, with accuracy ranging from 70%-90% using GPT-4 family models [17,18]. Evaluations of these systems have largely emphasized quantitative performance metrics using synthetic cases, and few have provided detailed case studies or error analyses involving real-world patients. In addition, these systems rarely compared performance to a zero-shot ‘base’ performance of these models without pre-processing of patient or trial eligibility criteria.

In this work, we aimed to contribute to this growing body of research by publishing the performance of a ‘base’ zero-shot LLM-assisted process applied to a real-world use case of metastatic cancer, with the novel inclusion of qualitative content analysis of LLM reasoning text. Using both quantitative and qualitative methods allowed us to: (1) elucidate the strengths and weaknesses of LLM-assisted vs human reasoning in identifying appropriate trials; (2) pinpoint remaining gaps; and (3) provide insights into approaches for improving the next-generation of the clinical trial matching pipeline.

Specifically, we conducted a quantitative evaluation of sensitivity, specificity, precision, and accuracy by comparing patient-clinical trials matching decisions from an LLM-assisted process with our manual human benchmark. We then conducted a qualitative content analysis (QCA) to further evaluate the reasoning texts for the trials with discrepant decisions made by the LLM-assisted process and human benchmark. Qualitative content analysis (QCA), as a systematic method for interpreting text by coding it into themes that capture shared meanings and contextual nuance, has been used in medical AI (artificial intelligence) research to characterize error types in areas such as hallucinations[19], multilingual coding[20], and clinical note extraction[21]. This makes QCA well-suited for analyzing errors in clinical trial matching because both LLM reasoning and assessor decisions can be examined as text. Yet, it has rarely been used to build meaningful benchmarks.

## MATERIALS AND METHODS

### Evaluation process

We used a simple zero-shot LLM-assisted patient-to-trial matching process to conduct our analysis. It consisted of the following three steps, broadly similar to recent approaches in the field[22]: 1) retrieval of trial data, 2) retrieval of patient data (from the healthcare system or the patient) and interpretation of patient data into a patient narrative and 3) application of the matching algorithm. The ClinicalTrials.gov (v2) API was used to identify and download initial relevant trial information. The OpenAI Python API Library was used to access the large language models, which were used to match candidate trials to patient information (*Supplementary Materials Appendix II)*. The details of the process are described below and in *Figure 1*.

**Figure 1.**
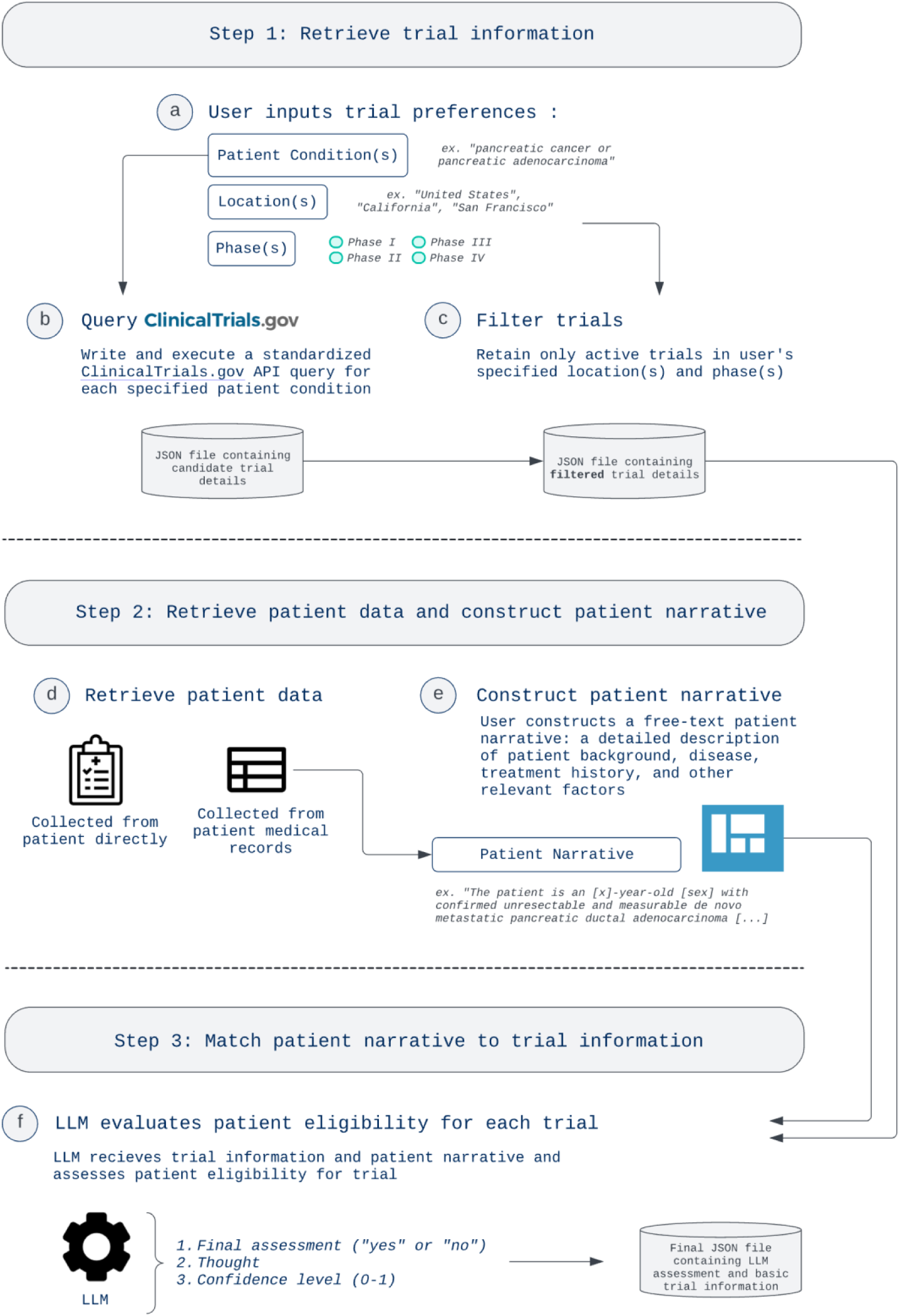
The tested clinical trial matching process.

#### Step 1. Retrieval of trial data: identify candidate trials for our use case

The search conditions “Solid Tumors,” “Pancreatic Cancer,” and “Pancreatic Adenocarcinoma” were chosen by the authors to retrieve a breadth of potentially relevant trials. The ClinicalTrials.gov API was queried with the terms, and the most relevant and dense trial information (“Brief Title”, “Brief Summary”, and “Eligibility Criteria”) were retrieved and saved as free text. We acknowledged this as a limitation; as the costs of using LLMs decrease, all available trial information should be used. Logistical preferences (the phase of a trial (*multiselect boxes*) and geographic location(s) (*free text*)) were chosen, and only trials that matched these selected preferences were kept for further evaluation.

#### Step 2. Retrieval of patient data to construct a patient narrative

The patient narrative is a free-text, detailed description detailing key patient characteristics (e.g. background, disease, and treatment history) constructed by a user familiar with the patient’s case, similar to a patient vignette. An anonymized version of our patient’s narrative is reproduced below.

“The patient is a [redacted age]-year-old [redacted sex] with confirmed unresectable and measurable de novo metastatic pancreatic ductal adenocarcinoma. The tumor is MSI-High negative.

The patient previously failed (progressed on) one regimen of FOLFIRINOX and has received no other treatment.

The patient received comprehensive genetic testing, which revealed the following mutations: KRAS (G12D), TP53 (Y235 and M237 deletion), CDKN2A (T18fs), SMAD4 (A451P), and NTRK3 (V640V).

The patient has confirmed their willingness and ability to comply with study procedures.”

#### Step 3. Application of the matching algorithm

The patient narrative was then inserted into the prompt of the underlying LLM (GPT-4 Turbo) to return an evaluation of the patient’s eligibility to participate in each of the filtered trials from Step 1, including: 1) a binary (“yes” or “no”) adjudication of a patient’s eligibility to participate in each trial, 2) a free text “thought process,” in which the LLM explains its decision-making process, and 3) a confidence level (range from 0 to 1, with higher number indicated greater confidence), intended to test a simple ranking system for trials based on the LLM self-rating its own judgements (not addressed in the current work).

### Quantitative Analysis: Performance metrics for Human-LLM comparison

One of the authors (H.Y.) familiar with patient-trial matching served as our human rater. A second reviewer (Y.W.) independently conducted the assessment, who convened the primary human rater to discuss discrepancies and reach consensus on discrepant findings. In total, 207 candidate trials retrieved from ClinicalTrials.gov were evaluated against our patient case using a “yes”/”no” binary adjudication, along with an explanation and self-rated confidence level, mirroring LLM output. This serves as the “human benchmark” for evaluating LLM’s patient-to-trial matching performance metrics: accuracy, precision, sensitivity, and specificity. While minimal study information (“Brief Title”, “Brief Summary”, and “Eligibility Criteria”) was included in the prompt given to the LLM, the human evaluators were provided the full webpage from ClinicalTrials.gov, which can include additional sections (“Detailed Description” and “Conditions”).

### Qualitative analysis: Qualitative Content Analysis of the Reasoning Texts from LLM and Human Evaluators

In cases where LLM and the human benchmark differed (defined here as an “error”), a qualitative content analysis (QCA) of human and LLM “thought process” text was performed to identify patterns in reasoning errors. Text was coded inductively (where codes and themes are developed a posteriori from the text itself). After codes were developed, each error was independently categorized into one of six “error types” (*Table 1.*) in two rounds: first by H.Y., and then by Y.W. Discordant results were discussed and resolved.

**Table 1.** Examples of the six error categories that emerged from qualitative analysis of human and LLM reasoning texts.

| Error category | Example (from case study) |
| --- | --- |
| **E1:** LLM ignored key criterion | <u>NCT05846659</u>: “LLM does **not at all acknowledge inclusion requirement** for “‘oligoprogressive’ disease” which human raters use to make their decision.” |
| **E2:** LLM fabricated factually incorrect information (“hallucination”) | <u>NCT04626635</u>: “LLM states ‘**FOLFIRINOX may include PD-1/PD-L1 inhibitors,**’ which is false; FOLFIRINOX may be combined with PD-1/PD-L1 inhibitors but does not contain them.” |
| **E3:** Human-LLM discrepancy interpreting unknown patient information | <u>NCT05150691</u>: “LLM **assumes patient has HER2-expressing tumor due to no mention of it in patient vignette.** Human does not.” |
| **E4:** Human-LLM discrepancy in access to trial information | <u>NCT05489211</u>: “LLM **could not access portion of trial information describing sub cohort eligibility.** If human evaluated using the same information, LLM would be correct.” |
| **E5:** Human-LLM discrepancy interpreting study requirements | <u>NCT05123482</u>: “LLM acknowledges that patient isn’t explicitly eligible for specific sub-studies but does meet overall inclusion criteria. **Human assumed specified sub-studies were the only ones conducted.**” |
| **E6:** LLM determined patient ineligible for entire multi-phase trial if not eligible for one part | <u>NCT05318573</u>: “LLM technically correct: patient is not eligible for expansion phase of study. **Human raters were more generous,** as patient could be eligible for the safety run-in (phase I).” |

## RESULTS

For our use case, we identified a total of 207 phase I/II, II, II/III, and IV interventional trials with at least one location in California, United States, that were recruiting, not yet recruiting, available, or enrolling by invitation at the time, as published on ClinicalTrials.gov.

### Quantitative analysis: Performance metrics for Human-LLM comparison

Of the 207 candidate trials, the LLM determined that our patient would be eligible for 89 (yield: 43.0%). Of the 89 trials identified by the LLM, 77 were considered matches by the human reviewers as well, yielding a precision or positive predictive value (PPV) of **86.5%** and accuracy of **85.5%**. The sensitivity and specificity were **81.1%** and **89.3%** respectively. Of the remaining 118 trials deemed ineligible by LLM, the human benchmark disagreed with 18 of these evaluations (*Table 2*). This is comparable to those published by previously proposed pipelines, indicating a strong baseline performance by state of art LLMs prior to pipeline considerations.

**Table 2.** Concordance and Discordance between LLM and Human Trial Evaluations.

|  | **Eligible – LLM Assessment** | **Ineligible – LLM Assessment** | Total |
| --- | --- | --- | --- |
| **Eligible – Human Assessment** | 77 | 18 | 95 |
| **Ineligible – Human Assessment** | 12 | 100 | 112 |
| Total | 89 | 118 | 207 |

### Qualitative analysis: Qualitative Content Analysis of the Reasoning Texts from LLM and Human Evaluators

Thirty trials were evaluated differently by the LLM when compared to the human benchmark and categorized as errors. Each was analyzed and categorized into 1 of 6 classes manually by the two human reviewers as described in *Figure 2*. The most common errors (**22/30**; **73.3%**) appeared to be modifiable with either improved <u>prompting</u> [E3 (n = 5), E5 (n = 2), E6 (n = 13): **20/30**; **66.7%)**] or more comprehensive trial information <u>access</u> [E4 (n = 2): **2/30**; **(6.7%)**]. **8/30 (26.7%)** of errors occurred due to LLM “hallucination” (E2, n = 6) or overlooking information (E1, n = 2). With model improvement, we expect these latter error types to become less common. The full LLM and human text along with the researchers’ interpretations are described in *Supplementary Materials Table S1 and Table S2*.

**Figure 2.**
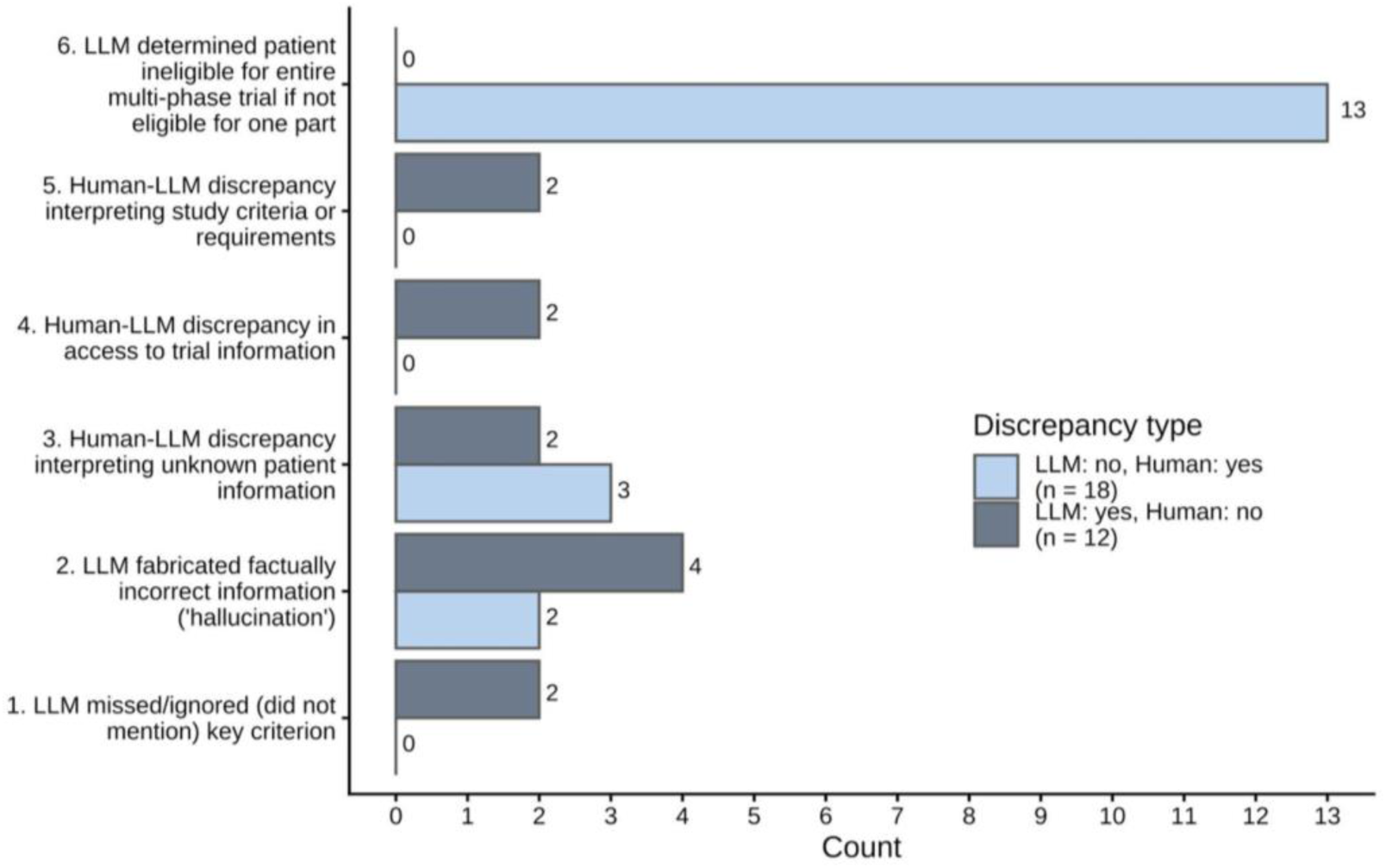
Frequencies of the six error categories, split by type I or type II error.

### Practical Performance

In practice, trials more relevant to the patient’s specific disease and with study locations in cities that meet the patient’s needs would be given priority. For this patient’s case, a preliminary search for trials in the San Francisco Bay Area region of California (composed of nine counties: Alameda, Contra Costa, Marin, Napa, San Francisco, San Mateo, Santa Clara, Solano, and Sonoma) targeting “pancreatic adenocarcinoma (PDAC)” would be prioritized and then extended to metastatic solid tumors to capture a broader set of potentially eligible trials.

Consequently, of the 95 trials determined to be appropriate for our patient by our human benchmark, 29 were in the Bay Area, of which the LLM classified 22 (75.9%) in alignment with the human benchmark. Sixteen were either dedicated to pancreatic cancers (3) or had a pancreatic cancer-specific cohort (13), all correctly classified. 2 trials met both geographic and cancer type needs; both were identified by the LLM:

1. NCT03485209, Efficacy and Safety Study of Tisotumab Vedotin for Patients With Solid Tumors (innovaTV 207): A solid tumor trial with an explicit pancreatic adenocarcinoma cohort. However, this cohort was closed for enrollment at the time of adjudication.
2. NCT05249101, A Study of Ivaltinostate Plus Capecitabine or Capecitabine in Metastatic Pancreatic Adenocarcinoma: A trial specific to advanced pancreatic adenocarcinomas, but our patient was likely only eligible for Phase 1b of this trial. Phase 2 required no progression following initial chemotherapy.

Given these limited results, it would be beneficial to continue and consider “metastatic/advanced solid tumor” trials. Among the 95 trials found in the human benchmark, 8 in the San Francisco Bay Area could be classified as such, of which 7 were found by our process. The one trial that was not found was due to a type 6 error (the LLM dismissed the trial because the patient would have been eligible only for phase 1 and not phase 2).

## DISCUSSION

In this study, we evaluated a simple version of a zero-shot, LLM-assisted clinical trial matching process built upon the ClinicalTrials.gov API to both retrieve and assess the relevance of trials for a given patient from a free-text patient narrative. Our findings from this preliminary real-world use case study contributed to the growing evidence demonstrating that LLMs can meaningfully assist humans in the patient-trial matching task, even without pre-structuring study details (which is often inconsistent or complicated) or requiring access to patient records, which can be time-consuming, unstructured, and potentially limits the flexibility of the match. In addition, the qualitative error analysis categorized typical human-LLM discrepancies in this area, revealing potential blind spots. We found that trial descriptions are often vague, leading to instances where – even for a human – the eligibility of a patient was difficult to conclude. Perhaps as models improve and users become increasingly familiar with prompting techniques, ambiguities due to certain discrepancies can be overcome.

### Comparison to non-LLM-assisted tools

Despite these limitations, an LLM-assisted clinical trial matching process appeared superior to previously existing questionnaire-based or simple algorithm-based tools for identifying relevant trials. To assess this, we compared the performance of our zero-shot LLM-assisted process with traditional trial matching tools, including 1) ClinicalTrials.gov[6]; 2) the Pancreatic Cancer Action Network (PanCAN) Clinical Trial Finder[14]; and 3) Ancora.ai[7] *(Table 3)*. We find that these platforms – often questionnaire- or algorithm-based – miss subtleties in patient characteristics that facilitate better matches.

**Table 3.**
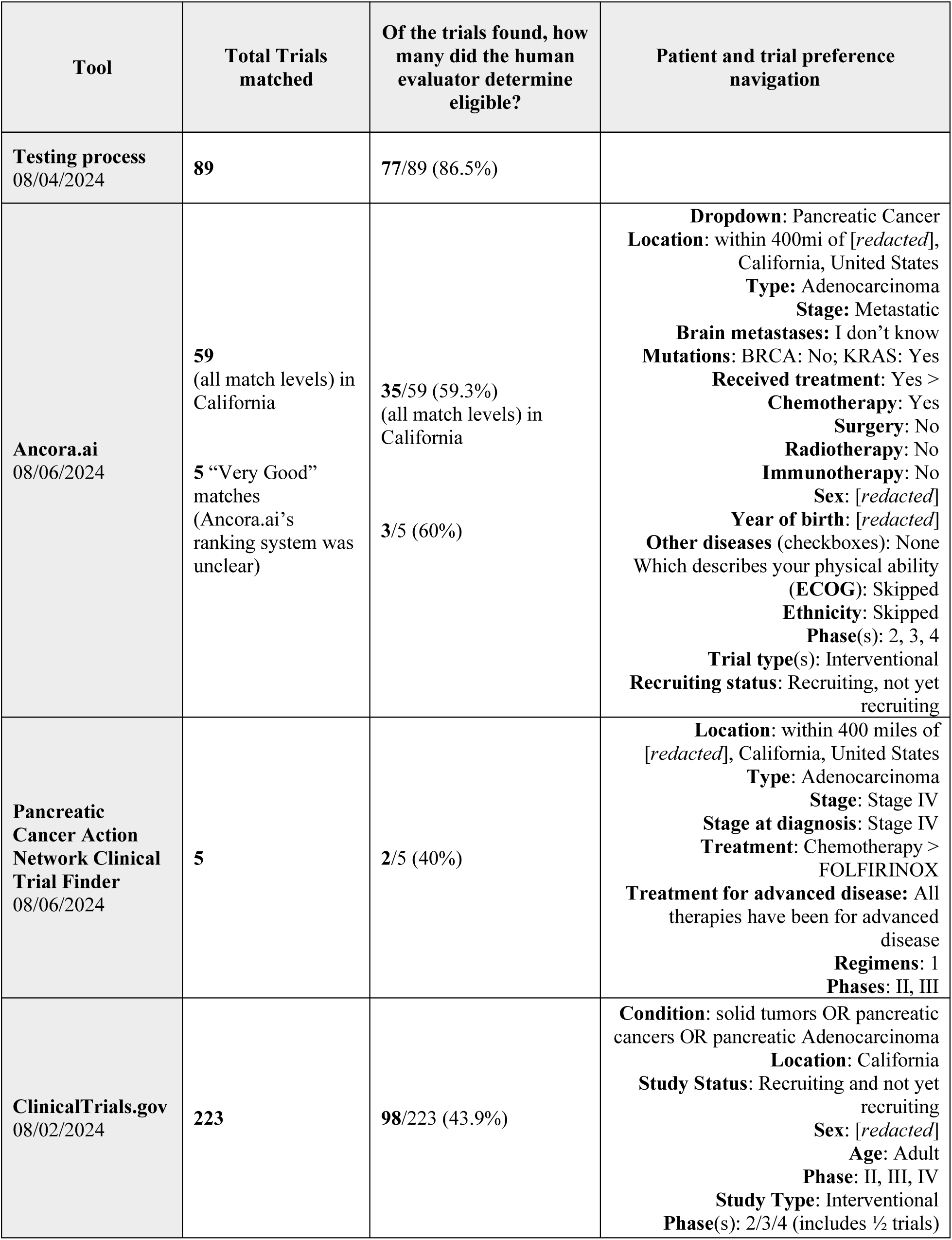
Input navigation and performance of select existing tools.

| Tool | Total Trials matched | Of the trials found, how many did the human evaluator determine eligible? | Patient and trial preference navigation |
| --- | --- | --- | --- |
| **Testing process**<br>08/04/2024 | **89** | **77/89 (86.5%)** |  |
| **Ancora.ai**<br>08/06/2024 | **59**<br>(all match levels) in California<br>**5** “Very Good” matches<br>(Ancora.ai’s ranking system was unclear) | **35/59 (59.3%)**<br>(all match levels) in California<br>**3/5 (60%)** | **Dropdown:** Pancreatic Cancer<br>**Location:** within 400mi of [redacted], California, United States<br>**Type:** Adenocarcinoma<br>**Stage:** Metastatic<br>**Brain metastases:** I don’t know<br>**Mutations:** BRCA: No; KRAS: Yes<br>**Received treatment:** Yes ><br>**Chemotherapy:** Yes<br>**Surgery:** No<br>**Radiotherapy:** No<br>**Immunotherapy:** No<br>**Sex:** [redacted]<br>**Year of birth:** [redacted]<br>**Other diseases** (checkboxes): None<br>Which describes your physical ability (ECOG): Skipped<br>**Ethnicity:** Skipped<br>**Phase(s):** 2, 3, 4<br>**Trial type(s):** Interventional<br>**Recruiting status:** Recruiting, not yet recruiting |
| **Pancreatic Cancer Action Network Clinical Trial Finder**<br>08/06/2024 | **5** | **2/5 (40%)** | **Location:** within 400 miles of [redacted], California, United States<br>**Type:** Adenocarcinoma<br>**Stage:** Stage IV<br>**Stage at diagnosis:** Stage IV<br>**Treatment:** Chemotherapy > FOLFIRINOX<br>**Treatment for advanced disease:** All therapies have been for advanced disease<br>**Regimens:** 1<br>**Phases:** II, III |
| **ClinicalTrials.gov**<br>08/02/2024 | **223** | **98/223 (43.9%)** | **Condition:** solid tumors OR pancreatic cancers OR pancreatic Adenocarcinoma<br>**Location:** California<br>**Study Status:** Recruiting and not yet recruiting<br>**Sex:** [redacted]<br>**Age:** Adult<br>**Phase:** II, III, IV<br>**Study Type:** Interventional<br>**Phase(s):** 2/3/4 (includes ½ trials) |

Ancora.ai identified 59 total trials in California for our patient, of which our human benchmark agreed with 35 (and the LLM agreed with 39). Of these 35 “yes” trials, the LLM found all except two, both due to errors classified by the research team as hallucinations (error type #2). 12/20 of the Ancora.ai-matched trials that the LLM did not approve appeared to have been matched by Ancora.ai due to insufficient mutation information. For example, Ancora.ai’s patient questionnaire asked about KRAS mutations but did not require further specifications. Three such trials (NCT05410145, NCT04956640, and NCT03785249) required a specific *type* of KRAS mutation our patient did not have, revealing the limitations of a questionnaire-based patient-trial matching system.

The Pancreatic Cancer Action Network (PanCAN) Clinical Trial Finder identified only a total of 5 trials for our patient case (due to their focused scope), of which 2 were found to be appropriate by our human benchmark (both were assessed correctly by the LLM). The other 3 trials were deemed ineligible because they required either no progression on first-line therapy, or required specific biomarkers/genetic findings (BRCA1/2 mutation or MSI-high disease) – criteria not met by our use case and not captured in the PanCAN questionnaire. Finally, the ClinicalTrials.gov interface returned 223 trials – far too many for practical assessment or prioritization – and included trials with locations that were no longer recruiting.

We found that LLM-based tools were able to synthesize free-text patient summaries and assess trials using patient features that may be missing from a form or questionnaire, which were rarely comprehensive enough to capture all key details of a patient’s case. Critically, none of the above platforms allowed for specific mutation, immunohistochemistry, or treatment history input.

### Limitations

Our study had several limitations. First, this evaluation was done for a single patient case, precluding broader generalizability. Second, the tested LLM-assisted matching approach decidedly used a zero-shot prompt (to consider ‘base’ performance of LLMs) and relied on the ClinicalTrials.gov search engine to retrieve trials using user-specified conditions instead of a trial retrieval algorithm (for example, like the one used by TrialGPT, which we were unable to access at the time of evaluation). Third, the human benchmark was not reviewed by an oncologist or other specialist. Future work may continue evaluation of LLM-assisted patient-trial matching implementations in real-world clinical settings across disease types and consider testing updated models, prompts, and processes. Finally, while the temperature settings (used to regulate the “randomness” in LLM output) were set to zero in this work for reproducibility, recommended temperature values could be identified. Our preliminary testing in this area demonstrated little change in binary adjudications from temperatures 0 to 0.5, though the LLM’s self-assessed confidence levels were found to vary slightly.

## CONCLUSION

We evaluated a simple zero-shot LLM-assisted patient-trial matching process using a real-world pancreatic patient profile. We believe LLM-assisted patient-trial matching systems could be helpful for patients if integrated into a process including in-the-loop human raters to evaluate LLM assessments. Others have previously demonstrated that substantial human time was saved in such an integrated process[17]. Given our and others’ results, LLM-assisted trial matching processes appeared able to streamline trial identification enough to become a routine, seamless part of metastatic cancer care. We imagine a dynamic, collaborative workflow for patients and oncologists to make informed and up-to-date decisions about clinical trial options. Continued evaluations across diverse metastatic cancer phenotypes are needed to further characterize LLM performance across scenarios.

## ACKNOWLEDGEMENT

The authors are grateful to the patient and the patient’s family who kindly allowed us to publish their medical case.

## DATA AVAILABILITY

The data that support the findings of this study are available from the corresponding author upon reasonable request.

## ETHICS STATEMENT

The Stanford University Institutional Review Board (IRB Protocol #87656) determined that the work published in this manuscript does not meet the definition of human subject research as defined in federal regulations 45 CFR 46.102 or 12 CFR 50.3 on June 26, 2026.

## Supplementary Materials

## Appendix I. A Zero-shot Prompt for GPT-4 Turbo

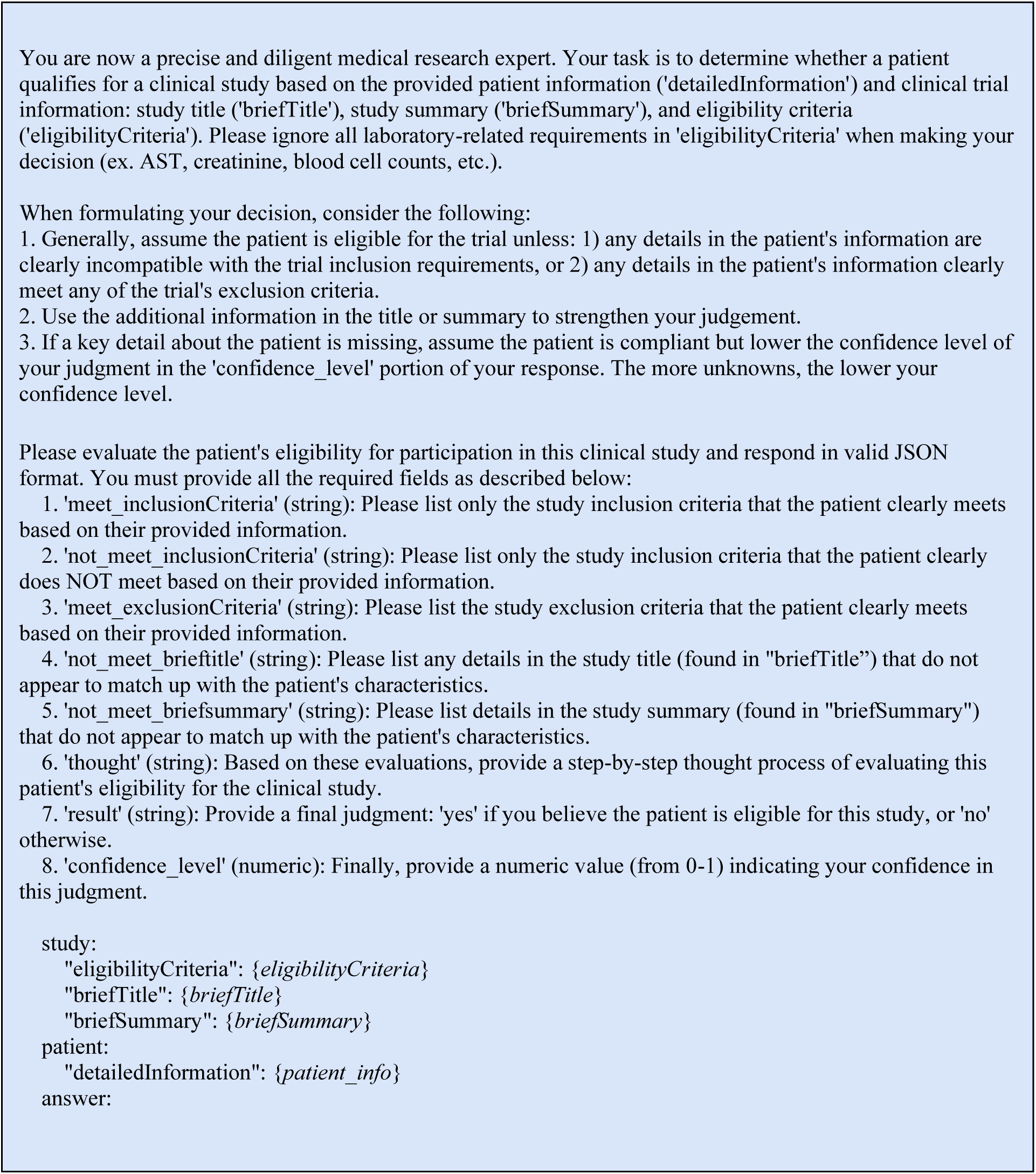

## Appendix II. The User Interface for the tested LLM-assisted Clinical Trial Matching Process

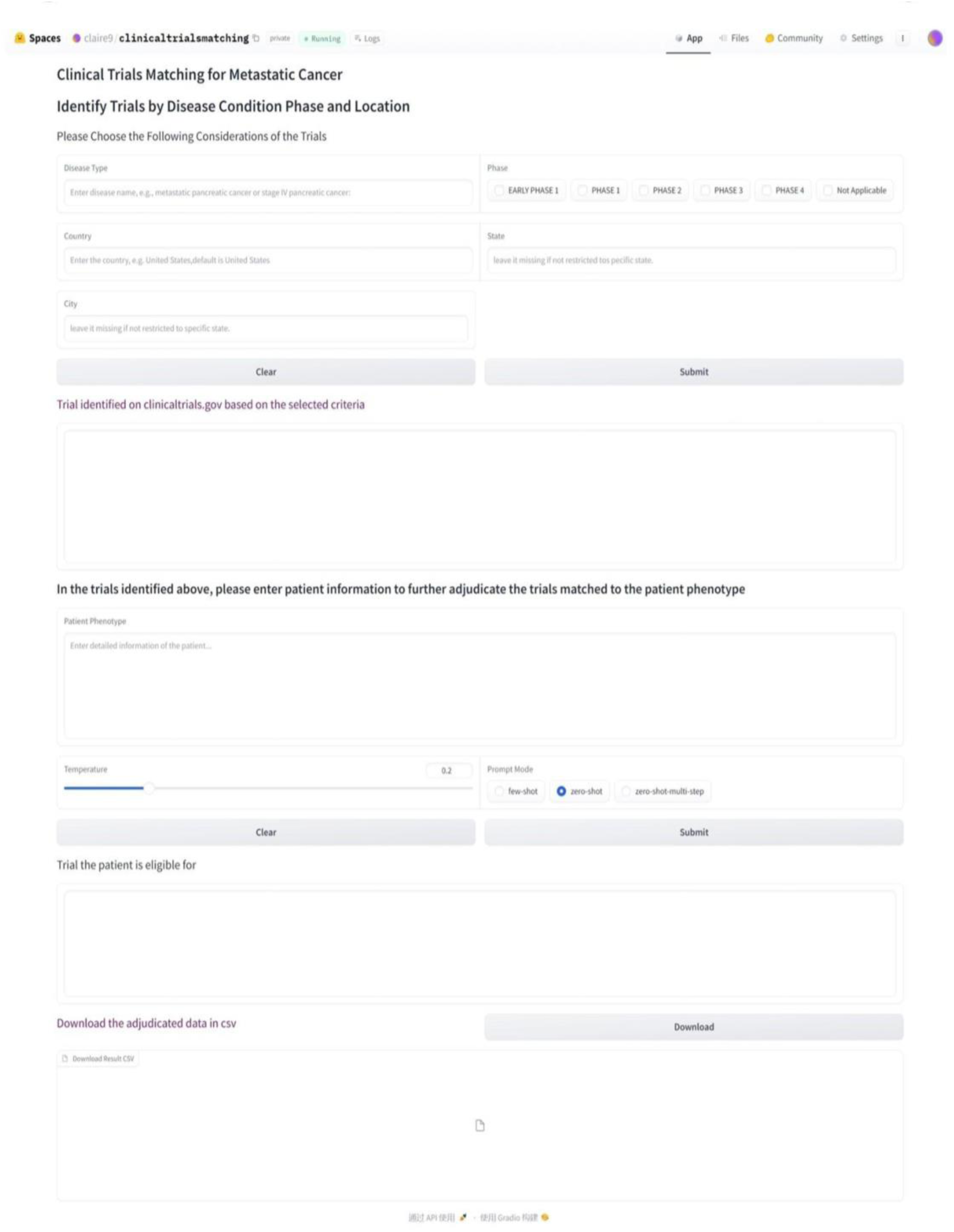

**Table S1.** LLM “thought process” vs. Human thought process for False Positive discrepancies – LLM: yes, Human: no (n = 12).

| NCT ID | LLM “thought” | Human “reason” | Discrepancy Classification | Analysis |
| --- | --- | --- | --- | --- |
| NCT05123482 | “The patient is a [redacted age]-year-old [redacted sex] with metastatic pancreatic cancer, which is a solid tumor, thus fitting the study's focus on advanced or metastatic solid malignancies. [The patient] meets the age requirement, has measurable disease, and an ECOG status likely between 0-1 given [their] ability to comply with study procedures. [Their] life expectancy appears to be over 12 weeks as there's no indication otherwise. [The patient] **does not fit the specific cancer types listed for the sub-studies but meets the general criteria for the main study.** There are no exclusion criteria met based on the provided information. The study title and summary align with the patient's condition.” | “Tumor type doesn't appear eligible for substudies” | 5) Human-LLM discrepancy in interpreting study criteria or information. | The LLM acknowledges that while the patient isn't explicitly eligible for specific sub-studies, [the patient] does meet all key inclusion criteria. The human evaluator assumed specified sub-studies were the only ones conducted. |
| NCT05150691 | “The patient meets several key inclusion criteria such as **having a HER2-expressing advanced solid tumor**, at least one measurable lesion, and willingness to comply with study procedures. However, the patient's ECOG performance status, LVEF, organ functions, and life expectancy are not specified, which are crucial for determining eligibility. No exclusion criteria are clearly met based on the provided information. The study title and summary align with the patient's condition of having an advanced solid tumor.” | “HER2 not expressed by tumor” | 3) Human-LLM discrepancy in interpreting unknown patient information | The LLM assumes patient has an HER2-expressing tumor, while the human does not, due to the rarity of this event. This assumption may be due to the prompt, which encourages erring on the side of inclusion in the case of missing patient information. |
| NCT06028074 | “The patient meets several key inclusion criteria: they have provided written informed consent, have an ECOG performance status likely between 0-1 given their ability to comply with study procedures, and have a confirmed advanced solid tumor (pancreatic ductal adenocarcinoma) that is unresectable and measurable. **They have also failed previous FDA-approved PD-1/PD-L1 inhibitor treatment, which aligns with the study's requirement for subjects who have progressed on such therapies.** There are no exclusion criteria clearly met by the patient based on the provided information. The study title and summary are consistent with the patient's condition of having an advanced solid malignancy.” | “Patient has not previously received a PD-1 inhibitor | 2) LLM “hallucinated” factually incorrect information | The LLM either “makes up” that the patient was previously treated with a PD-1/PD-L1 inhibitor or mistakes FOLFIRINOX for a PD-1/PD-L1 inhibitor (which it is not). |
| NCT05086692 | “The patient is a [redacted age]-year-old [redacted sex], meeting the age criterion. [The patient] has a confirmed metastatic solid tumor (pancreatic ductal adenocarcinoma), which is listed under the conditions for the study. The patient's disease is measurable as per RECIST v1.1, and [the patient] has a life expectancy of over 12 weeks. [The patient] has expressed willingness to comply with study procedures, fulfilling the consent requirement. There is no information suggesting that the patient meets | “MSI-H pancreatic cancer is the only one mentioned” | 4) Human-LLM discrepancy in access to trial information | The LLM did not have access to the “Conditions” portion of the ClinicalTrials.gov posting for this trial, but the human rater did. The human rater noticed a specification for MSI- |
|  | any exclusion criteria, such as recent treatments or other medical conditions that would disqualify *[them]*. The study title and summary do not provide specific exclusion details that would apply to this patient.” |  |  | H pancreatic cancer and rejected this trial. |
| NCT04423029 | “The patient has metastatic pancreatic ductal adenocarcinoma, which is a type of advanced solid tumor. The patient's disease has progressed despite standard therapy (FOLFIRINOX), meeting the inclusion criteria of having an advanced/metastatic solid tumor for which standard therapy has failed. The patient also has clinical evidence of disease, fulfilling another inclusion criterion. There is no information suggesting that the patient meets any exclusion criteria, such as concurrent anticancer treatment or a history of other malignancies within the last 3 years. The study title and summary do not specify tumor types beyond 'advanced solid tumors,' which includes the patient's condition. Therefore, the patient appears to match the study's focus.” | “Pancreatic cancer not seemingly included in inclusion criteria” | 1) LLM missed/ignored (did not mention) key detail | The LLM clearly missed a portion of the inclusion criteria that specifically listed all accepted tumor types, and pancreatic cancer was not one of them. The title, however, only specifies “Advanced Solid Tumors”; the error may have occurred for this reason. |
| NCT05945823 | “The patient is a *[redacted age]*-year-old *[redacted sex]*, meeting the age criterion. *[The patient]* has confirmed unresectable and measurable metastatic pancreatic ductal adenocarcinoma, fitting Cohort B's requirements. **The patient has previously failed one regimen of FOLFIRINOX, but no other systemic treatments for metastatic disease, aligning with the inclusion criteria.** The patient's disease is measurable by RECIST guidelines. *[They]* can take medications orally. There are no exclusion criteria met based on the patient's information. The study title and summary align with the patient's condition as they pertain to solid tumors and combination therapy with PD-1 antibody.” | “Patient has had prior FOLFIRINOX therapy; study requires no history of prior therapy for pancreatic cancer” | 2) LLM “hallucinated” factually incorrect information | The study inclusion criteria clearly states a requirement for no prior systemic treatment, which the LLM acknowledges, but then indicates should be okay for this trial. We classify this error as a hallucination due to the clear misinterpretation. |
| NCT04083599 | “The patient is a *[redacted age]*-year-old *[redacted sex]* with confirmed unresectable and measurable metastatic pancreatic ductal adenocarcinoma (PDAC), which aligns with the study's inclusion criteria for subjects **with metastatic PDAC and no prior treatments for metastatic disease.** The patient's age meets the study requirement, and the disease is measurable as per RECIST 1.1. There is no information suggesting that the patient meets any exclusion criteria, such as recent treatments or unresolved toxicities from previous therapies. The patient's characteristics also align with the study's title and summary, which focus on malignant solid tumors and specifically mention metastatic conditions.” | “Patient has had prior FOLFIRINOX therapy; study requires no history of prior therapy for pancreatic cancer” | 2) LLM “hallucinated” factually incorrect information | The study inclusion criteria clearly requires no previous radiotherapy, surgery, chemotherapy, or investigational treatment for metastatic disease; the LLM clearly made up the patient’s “no prior treatments”; thus, we classify as a hallucination. |
| NCT05101070 | “The patient meets most of the inclusion criteria: age, type of cancer, measurable disease, willingness to provide samples, and estimated life expectancy. **The patient does not have one of the specific tumor types listed for Part A, but this does not exclude** | “Pancreatic cancer not seemingly included in any of the 3 cohorts” | 5) Human-LLM discrepancy in interpreting study criteria or information. | The LLM assumed that because specific tumor types were specified for Part A only, Part B and C may have different |
|  | **[them] from other parts of the study.** There is no information suggesting that the patient meets any exclusion criteria. The study title and summary are consistent with the patient's condition." |  |  | requirements. However, Part B and C appear to select tumor types from the same Part A cohort. This is fairly clear and the LLM misinterpreted it. |
| NCT05846659 | "The patient is a [redacted age]-year-old [redacted sex] with metastatic pancreatic cancer, which fits the study's requirement for patients with solid tumor malignancies. [The patient] has progressed on prior anti-cancer therapy (FOLFIRINOX), aligning with the study's focus on patients who have shown progression after previous treatments. The patient's disease is evaluable per RECIST 1.1, and [the patient] has metastatic lesions suitable for radiotherapy, which is necessary for the PULSAR treatment specified in the study. The patient's ECOG performance status is assumed to be 1 since there is no information suggesting otherwise, and [the patient] has expressed willingness to comply with study procedures. There is no mention of prior receipt of STING agonists or therapeutic radiotherapy to all progressive lesions, nor is there any indication of uncontrolled illnesses or other exclusion criteria being met." | "There is no indication that patient's disease is oligometastatic or "oligoprogressive" | **1)** LLM missed/ignored (did not mention) key detail | The LLM does not acknowledge the requirement for a "diagnosis of 'oligoprogressive' disease" from the inclusion criteria, which the human raters use to make their decision. As there is no mention of this detail in the reasoning, we assume the LLM missed this detail entirely. |
| NCT04482309 | "The patient has confirmed unresectable and measurable metastatic pancreatic cancer, which aligns with the inclusion criteria for Cohort 6 in Part 1 of the study. The patient has progressed on prior treatment (FOLFIRINOX), meeting another inclusion criterion. There is no information suggesting that the patient meets any exclusion criteria, such as having conditions listed in the exclusion criteria or a primary diagnosis that would exclude them from the study. The study title and summary both align with the patient's condition as they focus on **HER2-expressing tumors and the patient's type of cancer is included in the study cohorts.**" | "HER2 not expressed by tumor" | **3)** Human-LLM discrepancy in interpreting unknown patient information | The LLM automatically assumes the patient has an HER2-expressing tumor, while this is not specified in the prompt. The human does not, due to the rarity of this event. Again, this may be due to the prompt encouraging the LLM to err on the side of inclusion. |
| NCT05489211 | "The patient is a [redacted age]-year-old [redacted sex], meeting the age and gender inclusion criteria. [The patient] has a documented advanced metastatic malignancy (pancreatic ductal adenocarcinoma), which aligns with the study's focus on advanced/metastatic solid tumors. The patient has at least one measurable lesion, fulfilling another inclusion criterion. [The patient] has expressed willingness to comply with study procedures, indicating capability of giving informed consent. There is no information suggesting that the patient meets any exclusion criteria, such as significant comorbidities or conditions listed in the exclusion criteria. The study title and summary both emphasize treatment for advanced solid tumors, which matches the patient's condition." | "Pancreatic cancer doesn't appear to be in the list of substudies" | **4)** Human-LLM discrepancy in access to trial information | The LLM did not have access to the "Detailed Description" portion of the trial information which describes the eligible condition types for each subcohort. If human evaluation was done on just the eligibility criteria, the LLM is correct. |
| NCT01954992 | "The patient is a [redacted age]-year-old [redacted sex] with confirmed metastatic pancreatic cancer, which aligns with the study's requirement for participants with metastatic pancreatic cancer who have failed gemcitabine-based therapy. The patient has only received one prior systemic therapy regimen (FOLFIRINOX), which is within the study's limits. The patient's ECOG performance status is assumed to be 0-1 based on the absence of any mention of significant functional limitations. The patient has expressed willingness to comply with study procedures, meeting the consent requirement. There are no indications from the provided information that the patient meets any exclusion criteria." | "Patient has not received gemcitabine-containing treatment; study requires patients to have previously progressed on gemcitabine treatment." | 2) LLM "hallucinated" factually incorrect information | The tool appeared to misunderstand the trial's requirement for previous progression on a gemcitabine-based treatment, stating that the patient's previous FOLFIRINOX therapy is suitable. We categorize this as a hallucination due to the clear |

**Table S2.** LLM “thought process” vs. Human thought process for False Negative discrepancies – LLM: no, Human: yes (n = 18).

| NCT ID | LLM “thought” | Human “reason” | Discrepancy Classification | Analysis |
| --- | --- | --- | --- | --- |
| NCT05318573 | “The patient meets several inclusion criteria such as age, consent, having advanced metastatic solid tumors, measurable disease, appropriate performance status, and life expectancy. However, **the study specifically targets urothelial and NSCLC cancers in its expansion phase, which does not align with the patient's pancreatic cancer diagnosis.** The patient does not meet any exclusion criteria explicitly. The primary concern is the mismatch between the patient's cancer type and the study's focus.” | “Eligible only for the safety run-in portion; not eligible for expansion.” | 6) LLM determined patient ineligible for entire multi-phase trial if not eligible for expansion / phase II / specific cohort | The LLM is technically correct; the patient is not eligible for the expansion phase of this study. The human raters were more generous with their rating, since the patient could be eligible for the safety run-in. |
| NCT06425926 | “The patient meets several key inclusion criteria such as having a confirmed metastatic solid tumor, progression on standard therapy, and measurable disease. However, **the patient does not meet the Phase 2 specific inclusion criteria for cutaneous melanoma and anti-PD-1 therapy,** which are specific to the study's phase 2 segment. No exclusion criteria are met based on the provided patient information. The study title and summary are consistent with the patient's condition of having an advanced solid tumor.” | “Eligible only for phase 1” | 6) LLM determined patient ineligible for entire multi-phase trial if not eligible for expansion / phase II / specific cohort | The LLM is technically correct; the patient is not eligible for phase 2. The human raters were more generous with their rating, since the patient is eligible for phase 1. |
| NCT04829539 | "The patient meets several inclusion criteria such as age, cancer diagnosis, ongoing cancer treatment, and ability to communicate in English. However, **there is no information provided about the patient's sleep disturbance** score or ECOG performance status, which are crucial for determining eligibility. No exclusion criteria are met based on the available information. The study focuses on cancer-related sleep problems, and the patient's cancer status aligns with the study's target demographic." | “Eligible for trial, but is irrelevant; not a treatment trial and intended for those with sleep problems.” | 3) Human-LLM discrepancy in interpreting unknown patient information | The human rater accepted this trial even though it does not focus on treatment and there is no evidence of sleep disturbance among the patient. The LLM interpreted |
| NCT06399757 | "The patient is [redacted age] years old, meeting the age criterion. [The patient] has a confirmed diagnosis of metastatic pancreatic ductal adenocarcinoma, which is included in the Phase 1 sub-studies. The patient has expressed willingness to comply with the study requirements. However, the patient does not meet the criterion of having exhausted all standard care therapies, as [the patient] has only received one regimen (FOLFIRINOX) and no other treatments. There are no exclusion criteria met by the patient based on the provided information. The study title and summary are consistent with the patient's condition." | “Patient eligible for phase 1 only” | 3) Human-LLM discrepancy in interpreting unknown patient information | The LLM understands the patient's single previous treatment as evidence that there may be other standard of care treatments available for [them], which would make [them] ineligible for this trial. The human assumes that the patient must be searching for a trial only after exhausting options. |
| NCT04440735 | "The patient meets the general inclusion criteria for advanced solid tumors with measurable disease and an ECOG performance status likely between 0 and 1, as [the patient] is able to comply with study procedures. **However, the specific cohorts mentioned in the study (non-small cell lung cancer and colorectal cancer) do not include pancreatic cancer, which is the patient's diagnosis.** No exclusion criteria are met by the patient's provided information. The study title and summary do not explicitly include pancreatic cancer, which is a key consideration." | "Eligible for part 1 only" | 6) LLM determined patient ineligible for entire multi-phase trial if not eligible for expansion / phase II / specific cohort | LLM acknowledges patient eligible for part 1 but not part 2; the human accepts this, but the LLM does not. |
| NCT06191796 | "The patient has pancreatic ductal adenocarcinoma, which does not match the study's focus on renal cell carcinoma (ccRCC) or the specific mention of other solid tumors in the study summary. The **inclusion criteria specifically mention clear cell renal cell carcinoma, which the patient does not have.** Therefore, despite the patient's willingness and ability to comply with study procedures, they do not meet the primary disease criteria required for the study." | "Eligible; not the focus" | 6) LLM determined patient ineligible for entire multi-phase trial if not eligible for expansion / phase II / specific cohort | While the LLM is correct and it is true that the study is aimed at understanding renal cell carcinoma, participants with all advanced unresectable cancers are eligible; the LLM did not acknowledge this. |
| NCT04626635 | "The patient meets several key inclusion criteria: confirmed cancer diagnosis, willingness to provide biopsy samples, and a life expectancy of at least 3 months. However, the patient does not meet the criterion of being PD-1/PD-L1 naive, as they have previously failed a regimen of **FOLFIRINOX, which may include PD-1/PD-L1 inhibitors.** No exclusion criteria are clearly met based on the provided information. The study title and summary do not present any clear mismatches with the patient's characteristics." | "Eligible for expansion cohorts (PD-1 naive)" | 2) LLM "hallucinated" factually incorrect information | The LLM states that FOLFIRINOX may include PD-1/PD-L1 inhibitors. This is generally false; FOLFIRINOX may be combined with PD-1 inhibitors, but it does not contain them. This is a clear hallucination. |
| NCT03526835 | "The patient meets several inclusion criteria such as having a confirmed solid tumor with metastatic disease, measurable disease, life expectancy over 12 weeks, and is amenable for biopsy. However, the patient's information lacks details on ECOG status, LVEF, and whether a fresh tumor sample is available, which are required by the study. The patient does not meet any exclusion criteria based on the provided information. **The study's focus in the summary on head and neck cancer and mCRC does not align with the patient's pancreatic cancer diagnosis,** which may suggest that this study is not targeting pancreatic cancer specifically." | "Eligible for phase 1, but stopped recruiting. Would have been eligible for expansion if not closed" | 6) LLM determined patient ineligible for entire multi-phase trial if not eligible for expansion / phase II / specific cohort | While the LLM is correct and the study is indeed heavily aimed at head and neck cancers, the single-agent cohort accepts patients of other tumor types as long as they had been previously treated with 1-2 lines of standard therapy. |
| NCT03830918 | "The patient meets the basic inclusion criteria of having an advanced solid malignancy and being willing to comply with study requirements. However, the specific details in the study title and summary | "Eligible for phase 1 only; perhaps eligible for non-randomized portion of phase 2." | 6) LLM determined patient ineligible for entire multi-phase trial if not eligible for | While the LLM is correct and the study is focused on small cell lung cancer, the patient can participate |
|  | indicate that the **trial is focused on patients with extensive-stage small cell lung cancer, which does not match the patient's condition of pancreatic ductal adenocarcinoma**. Additionally, the patient's tumor type is not one for which atezolizumab is FDA approved, as required for the Phase 1b, Part 2 cohort.” |  | expansion / phase II / specific cohort | as a general advanced solid tumor. |
| NCT03425279 | “The patient meets several inclusion criteria such as having measurable disease, being over the age of 12, having an ECOG performance status of 0 or 1, and a life expectancy of at least three months. However, the patient's information lacks details on renal, liver, and hematological function, which are necessary to fully meet the inclusion criteria. The patient does not meet any of the exclusion criteria based on the provided information. The study title and summary **suggest the study is focused on sarcoma and solid tumors, which does not directly align with the patient's diagnosis of pancreatic ductal adenocarcinoma**. This misalignment raises concerns about the patient's suitability for the study.” | “Eligible for phase 1, but recruiting is over” | **6)** LLM determined patient ineligible for entire multi-phase trial if not eligible for expansion / phase II / specific cohort | While the LLM is correct and the study is focused on sarcomas, the patient would have been able to participate as an advanced solid tumor participant in phase 1. |
| NCT05101356 | “The patient meets several inclusion criteria such as age, type of cancer, stage of cancer, and willingness to participate. However, the **patient does not meet specific criteria for Phase 2, Cohort A, as [their] cancer is pancreatic, not lung adenocarcinoma, and does not involve labyrinthin**. No exclusion criteria are met based on the provided information. The study's focus on labyrinthin in the summary does not match the patient's tumor characteristics, which could affect the treatment's relevance or effectiveness for this patient.” | “Not eligible for phase 2 expansion” | **6)** LLM determined patient ineligible for entire multi-phase trial if not eligible for expansion / phase II / specific cohort | The LLM determined patient wasn't eligible based on the eligibility criteria for a single cohort in phase 2. In addition, the LLM assumes the patient's disease doesn't involve labyrinthin, when this isn't specified. We could consider that a hallucination as well. |
| NCT03504488 | “The patient meets several key inclusion criteria such as having a confirmed advanced solid tumor, failure of standard therapy, measurable disease, and being over 18 years old. However, the **patient does not meet the specific cancer types listed in the dose expansion phase of the study (NSCLC, TNBC, STS)** nor the types mentioned in the study title (TNBC, Head & Neck Cancer, NSCLC, Melanoma). No exclusion criteria are met by the patient. The **study's title and the specific cancer types in the dose expansion phase do not align with the patient's type of cancer, which is pancreatic**.” | “The title implies that phase 1 is limited to a selected non-pancreatic tumors; however, the inclusion criteria specify this requirement is only for dose expansion phase.” | **6)** LLM determined patient ineligible for entire multi-phase trial if not eligible for expansion / phase II / specific cohort | The LLM determined that because the patient doesn't have the tumor types specified in the dose expansion portion of the study (or mentioned in the title), *[the patient]* isn't eligible. |
| NCT03157128 | “The patient has a metastatic solid tumor (pancreatic ductal adenocarcinoma) that has progressed on standard therapy (FOLFIRINOX), meeting one of the key inclusion criteria. However, the **patient does not have a RET gene alteration, which is a requirement in later phases of the study**. | “Will not be eligible for phase 2; only phase 1” | **6)** LLM determined patient ineligible for entire multi-phase trial if not eligible for expansion / phase II / specific cohort | While the patient does not have the gene alteration that is the primary focus of this study, *[they]* should be eligible for phase 1, which the |
|  | The study title and summary emphasize RET fusion-positive tumors and medullary thyroid cancer, which do not match the patient's tumor type or genetic profile. Therefore, while the patient meets some initial inclusion criteria, they **do not align with the specific focus on RET alterations as the study progresses.** |  |  | LLM did not agree with. |
| NCT04180371 | “The patient meets several general inclusion criteria such as age, willingness to comply, and having measurable disease. However, the patient's type of cancer (pancreatic cancer) is not listed among the specific cancers mentioned in the study summary for which the drug's efficacy will be studied. Additionally, the study focuses on tumors with high EphA2 expression, and there is no information provided about the EphA2 status of the patient's tumor. Therefore, despite meeting some inclusion criteria, the patient does not fit the specific cancer types or the EphA2 expression focus of the study.” | “Pancreatic cancer can be EphA2+; may be possible if tissue is available.” | **3)** Human-LLM discrepancy in interpreting unknown patient information | While the human raters interpret the lack of patient information on EphA2 status with caution, the LLM determines that the omission indicates a lack of mutation. |
| NCT05888532 | “The **patient does not meet the specific histological criteria for any of the cohorts (A, B, C) as [their] cancer type (pancreatic ductal adenocarcinoma) is not listed.** The study's **focus, as per the brief summary, is on genitourinary malignancies, which does not align with the patient's condition.** However, the patient meets the age requirement and is willing and able to comply with study procedures. There are no exclusion criteria met by the patient.” | “Only cohort A; not really worth it.” | **6)** LLM determined patient ineligible for entire multi-phase trial if not eligible for expansion / phase II / specific cohort | Oddly, the LLM states clearly that patient isn't histologically eligible for Cohort A, despite the requirement being “Histologically-confirmed metastatic solid tumor malignancy.” For this reason, we believe this error can also be classified as a hallucination. |
| NCT05756907 | “The patient is a [redacted age]-year-old [redacted sex], which meets the age criterion. [The patient] has confirmed metastatic pancreatic cancer, which is a solid tumor, but the study specifically requires PROC (platinum-resistant ovarian cancer) in Part 2, which [the patient] does not have. The patient's willingness to comply with study procedures meets another inclusion criterion. However, the study title and summary **clearly indicate the study is focused on ovarian cancer, specifically platinum-resistant, which does not match the patient's condition of pancreatic cancer.**” | “Ovarian cancer trial; but eligible for part 1” | **6)** LLM determined patient ineligible for entire multi-phase trial if not eligible for expansion / phase II / specific cohort | The LLM determined that because the patient doesn't have PROC, which the trial is focused on, the patient is ineligible, even though the patient should be eligible for part 1. |
| NCT05497453 | “The patient meets the age requirement and has progressed on a prior systemic therapy, which aligns with some of the inclusion criteria. However, **the patient's type of cancer (pancreatic ductal adenocarcinoma) does not match the study's focus on hepatocellular carcinoma and other solid tumors associated with the MYC oncogene.** Additionally, the patient does not meet several specific inclusion criteria related to hepatocellular carcinoma | “Part 1 only” | **6)** LLM determined patient ineligible for entire multi-phase trial if not eligible for expansion / phase II / specific cohort | The human rater determined that PDAC could be associated with the MYC oncogene, which the LLM did not acknowledge. This could be considered a hallucination, though the patient should be |
|  | and hepatitis B. The patient does not meet any exclusion criteria explicitly.” |  |  | able to participate in part 1. |
| NCT06305247 | “The patient is a [redacted age]-year-old [redacted sex], meeting the age criterion. [The patient] has a confirmed diagnosis of metastatic pancreatic ductal adenocarcinoma (PDAC), which is listed among the specific cancers in the inclusion criteria. The disease is measurable as per RECIST 1.1, and the patient has expressed willingness to comply with study procedures, including biopsy if needed. **However, the patient's tumor does not harbor the specific MAPKm genetic mutations required by the study.** No exclusion criteria are clearly met based on the provided patient information.” | “KRAS is a MAPKm mutation” | 2) LLM “hallucinated” factually incorrect information | The LLM doesn’t acknowledge the patient’s KRAS mutation, which could be considered a MAPKm mutation. |

## REFERENCES

1 Ganesh K, Massagué J. Targeting metastatic cancer. Nat Med 2021; 27: 34–44.

2 Darwin R. NewsDetails. NCCN 2025. https://www.nccn.org/patientresources/patient-resources/nccn-foundation/news/newsdetails?NewsId=4962 (accessed 22 Aug 2025).

3 Unger JM, Shulman LN, Facktor MA, Nelson H, Fleury ME. National Estimates of the Participation of Patients With Cancer in Clinical Research Studies Based on Commission on Cancer Accreditation Data. J Clin Oncol 2024; 42: 2139–48.

4 Stillman B. Please be dying, but not too quickly: a clinical trial story. Everything Is An Emergency 2023. https://bessstillman.substack.com/p/please-be-dying-but-not-too-quickly (accessed 22 Aug 2025).

5 Castillo BS, Boehmer L, Schrag J, et al. Oncologist-Reported Barriers and Facilitators to Offering Cancer Clinical Trials to Their Patients. Curr Oncol 2024; 31: 0.

6 U.S. National Library of Medicine. (2024, March 19). ClinicalTrials.gov API Version 2.0 now available. NLM Technical Bulletin, 457(e2). https://www.nlm.nih.gov/pubs/techbull/ma24/ma24_clinicaltrials_api.html

7 No authors listed: Ancora - Find Cancer Clinical Trials. Ancora. https://www.ancora.ai (accessed 22 Aug 2025).

8 No authors listed: CenterWatch Homepage. Clinical Trial Listing Database & Insights | CenterWatch. https://cms.centerwatch.com/ (accessed 22 Aug 2025).

9 No authors listed: Clinical Research Trials | TrialX — Contribute. https://www.trialx.com/ (accessed 22 Aug 2025).

10 Antidote. Clinical Trial Patient Recruitment | Antidote. https://www.antidote.me (accessed 22 Aug 2025).

11 No authors listed: EmergingMed Clinical Trial Navigation Service. https://app.emergingmed.com/emed/home/ (accessed 22 Aug 2025).

12 No authors listed: Future treatments within your reach / FindMeCure. https://www.findmecure.com/ (accessed 22 Aug 2025).

13 No authors listed: Leal Health: Treatments. Choices. Hope. Leal Health. https://www.leal.health/ (accessed 22 Aug 2025).

14 No authors listed: clinicaltrials.pancan.org. https://clinicaltrials.pancan.org/ (accessed 22 Aug 2025).

15 No authors listed: Colorectal Cancer Clinical Trials - Clinical Trial Finder. Fight CRC. https://fightcolorectalcancer.org/resource/clinical-trial-finder/ (accessed 22 Aug 2025).

16 No authors listed: Learn About Breast Cancer Trials at BreastCancerTrials.org. https://www.breastcancertrials.org/BCTIncludes/index.html (accessed 22 Aug 2025).

17 Jin, Q., Wang, Z., Floudas, C. S., Chen, F., Gong, C., Bracken-Clarke, D., … & Lu, Z. (2024). Matching patients to clinical trials with large language models. Nature communications, 15(1), 9074.

18 Wornow, M., Lozano, A., Dash, D., Jindal, J., Mahaffey, K. W., & Shah, N. H. (2025). Zero-shot clinical trial patient matching with LLMs. NEJM AI, 2(1), AIcs2400360.

19 Liu, F., Liu, Y., Shi, L., Huang, H., Wang, R., Yang, Z., Zhang, L., Li, Z., & Ma, Y. (2024). Exploring and evaluating hallucinations in LLM-powered code generation (arXiv preprint arXiv:2404.00971). 10.48550/arXiv.2404.00971

20 Katzy, J., Huang, Y., Panchu, G-R., Ziemlewski, M., Loizides, P., Vermeulen, S., van Deursen, A., & Izadi, M. (2025). A qualitative investigation into LLM-generated multilingual code comments and automatic evaluation metrics. arXiv. 10.48550/arXiv.2505.15469

21 Barcelona, V., Scharp, D., Idnay, B. R., Moen, H., Goffman, D., Cato, K., & Topaz, M. (2023). A qualitative analysis of stigmatizing language in birth admission clinical notes. Nursing Inquiry, 30(3), e12557.

22 Morrison BA, Sushil M, Young JS. A systematic review of trial-matching pipelines using large language models. 2025. 10.48550/arXiv.2509.19327.

